# Bayesian Networks in Healthcare: the chasm between research enthusiasm and clinical adoption

**DOI:** 10.1101/2020.06.04.20122911

**Authors:** Evangelia Kyrimi, Scott McLachlan, Kudakwashe Dube, Norman Fenton

**Author notes:** Corresponding author: Email address (Evangelia Kyrimi) Postal address: 10, Godward Square, Queen Mary University of London, Mile End Rd, Bethnal Green, London E1 4FZ.

## Abstract

**Problem:** Bayesian Networks (BN) can address real-world decision-making problems, and there is enormous and rapidly increasing interest in their use in healthcare. Yet, despite thousands of BNs in healthcare papers published yearly, evidence of their adoption in practice is extremely limited and there is no consensus on why.

**Method:** A preliminary review was conducted to identify research gaps and justify the conduct of a broader scoping or systematic review of BNs in healthcare

**Results:** We highlight that: (1) there have been no significant attempts to systematically review the domain; (2) there are weaknesses in the way BN development processes are presented in the literature; and (3) in contrast to enthusiasm (including from clinicians) there has been negligible adoption of published BNs into clinical practice.

**Conclusion:** A systematic review of BNs in healthcare is needed to a) understand the chasm between research enthusiasm and clinical adoption; and b) improve clinical adoption.

## 1. Introduction

Clinical decision making is a complex evolving process where evidence is gathered and diagnostic and treatment decisions are made (Tiffen, Corbridge and Slimmer, 2014). While clinicians are generally considered to be good decision-makers, it can be challenging to combine all available evidence and accurately reason under conditions of uncertainty (Bornstein and Emler, 2001). Many clinical decision support systems (CDSS) have been developed, and graphical probabilistic models such as Bayesian Networks (BNs) are one popular approach (Druzdzel and Flynn, 2002; Beattie and Nelson, 2006; Patel *et al*., 2009; Adams and Leveson, 2012; Shortliffe and Cimino, 2013; Lucas, 2001; Lucas, van der Gaag and Abu-Hanna, 2004).

BNs are based on Bayes’ theorem and model causal or influential relationships between variables. Bayes’ theorem represents a formal method for understanding the natural thought process people undergo while evolving their understanding of the likelihood of one phenomenon under observation as their knowledge of related influential phenomena increases. For instance, when clinicians perform differential diagnosis where multiple diseases present with similar symptoms: as new information from diagnostic tests and clinical examination of the patient become available, clinicians update their belief on what they believe is the illness causing the patient’s poor health. BNs use a graphical approach for *compact representation of multivariate probability distributions* and *efficient reasoning under uncertainty*. Their popularity in healthcare results from their interpretable graphical structure and their ability to: (i) model complex problems with causal dependencies where a significant degree of uncertainty is involved; (ii) combine different sources of information such as data and experts’ judgement; and (iii) model interventions and reason both diagnostically and prognostically. Based on limited marketing materials, there are indications that some major healthcare technology companies are using BNs in their non-public research and application development (Armstrong, 2018), including what is claimed as the largest BN in the world (MMC Ventures, 2017).

While working on several projects related to clinical decision support using BNs we realised that: (i) even though a vast number of healthcare related BNs are published every year, no existing paper describes the scope of BNs developed for use in healthcare; (ii) apart from a small number of micro-reviews about BNs for specific medical conditions, e.g. (Bielza and Larranaga, 2014) and one small-scale epidemiology-focused review on directed acyclic graphs that is yet to be peer reviewed (Tennant *et al*., 2019), the domain lacks a systematic or scoping review; and, (iii) that while some researchers have investigated potential reasons why a model might not be useful in practice, e.g. (Wyatt and Altman, 1995; Blackmore, 2005; Reilly and Evans, 2006; Moons *et al*., 2009; Adams and Leveson, 2012), no review investigates which of the published medical BNs identifies or acknowledges the reasons why a model may or may not have been adopted in healthcare.

In contrast to widely discussed and accepted benefits of BNs, it appears that negligible clinical adoption of *published* BNs into healthcare practice has occurred. We believe that it is only when we understand the necessary rules for adopting a BN in clinical practice, and whether or not the BNs described in the literature adhere to these rules, that we may bridge the gap between development and adoption of BNs in healthcare. *It is our intention in this work to draw attention to this gap through a small-scale investigation that briefly explores the challenges inhibiting BN development and adoption for healthcare application*.

### The remainder of this paper is organised as follows

Section 2 presents the method used to achieve the objectives of this paper; Section 3 discusses the results; Section 4 proposes the way forward; Section 5 presents our concluding remarks.

## 2. Method

A search using Queen Mary University’s library search system that includes access to PubMed, MedLine, ScienceDirect, Scopus, DOAJ and Elsevier was performed using a selection of keywords arranged in the following search algorithm:

~~~
“(((Bayes OR Bayesian) AND network) OR (probabilistic AND graphical AND model)) AND (medical OR clinical)”
~~~

To identify the existing knowledge gaps in this research area, a small-scale review was proposed to analyse a limited number of papers from which we could identify research gaps and develop an overarching view of the domain sufficient to justify effort for conduct of a broader scoping or systematic review of BNs in healthcare. Initially, 50 papers were selected based on their relevance to the search term as reported by the search engine. Relevance was computed based on both the number of times the search terms appear in the paper, and where within the paper the search terms appeared (title, abstract, keywords, keywords plus).

Table 1 identifies the basic review framework which was agreed by all of the authors of this paper. This framework was applied to the preliminary survey presented in this paper. It consists of a set of elements relating to generalisability of the development methodology and adoption of the BN in healthcare

**Table 1:** Preliminary Review Framework

|  |  |
| --- | --- |
| 1. *Generalisability of BN development process and its outcomes:* | ☐ Overall development process<br>☐ BN structure elicitation from knowledge<br>☐ BN parameters elicitation from knowledge<br>☐ BN structure learning from data<br>☐ BN parameters learning from data |
| 2. *BN adoption in healthcare:* | ☐ Clinical benefit<br>☐ Clinical credibility<br>☐ Accuracy<br>☐ Generalisability<br>☐ Usability<br>☐ Potential impact |
|  | Actual impact<br>In use |

## 3. Results and Discussion

### Literature on BNs in Healthcare

Figure 1 demonstrates the year on year increase of publications on BNs in healthcare both in total and when narrowed to identify *the most highly relevant* ones.

**Figure 1:**
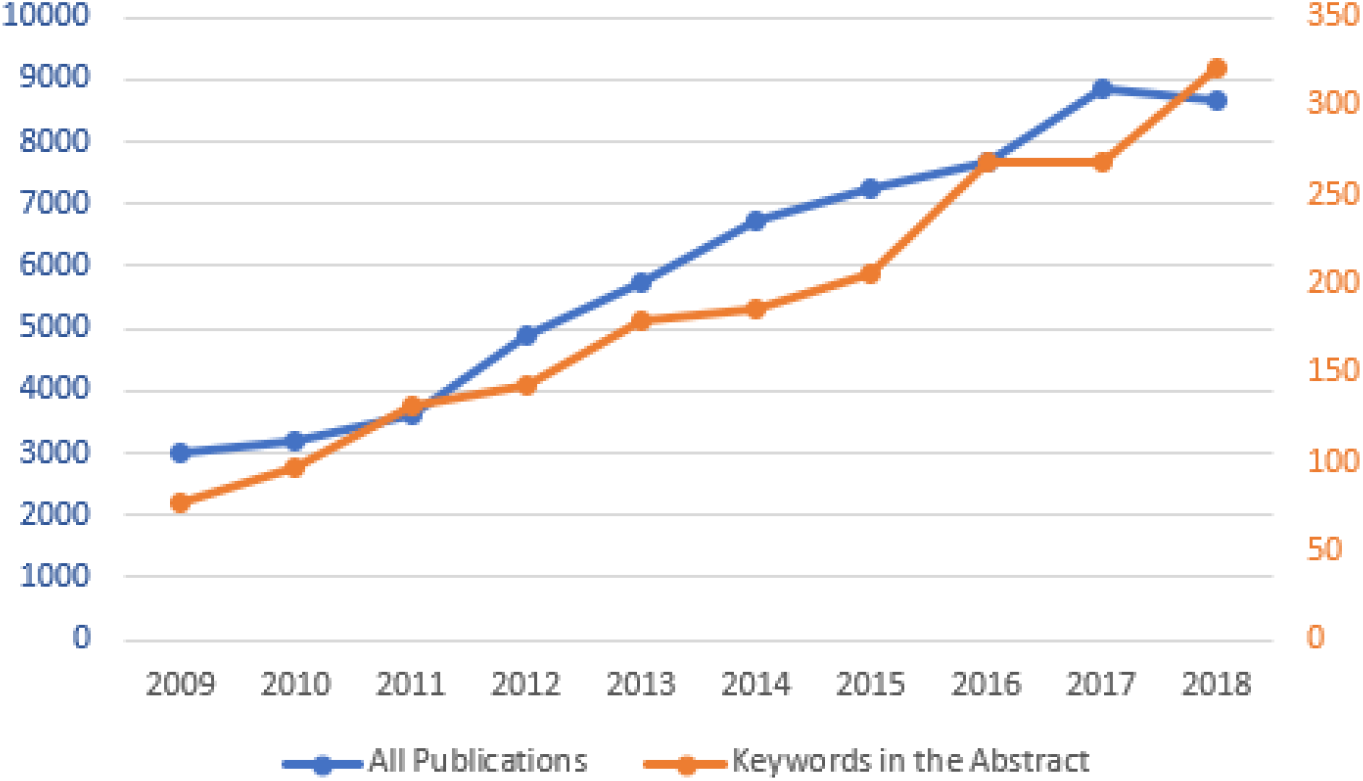
Number of publications on medical Bayesian networks per year

### Application of Preliminary Review Framework

Our objectives were to investigate: (1) generalisability of the BN development process; and, (2) BN adoption in healthcare. For inclusion, all authors agreed that papers had to possess all of the following criteria:

1. Describe a genuine BN model (or BN modelling process) rather than Bayesian statistics or naive Bayes;
2. Be targeted clearly at a medical problem; and,
3. Be intended to support clinicians or patients in decision making or prediction.

Our initial search identified a collection of 1894 papers for screening. Screening involved excluding papers: (i) that were duplicates; (ii) published outside the period 1995–2018; (iii) not published in English; (iv) not published in journal or conference proceedings; and (v) whose primary content was not healthcare related. After screening, 834 papers remained for eligibility assessment. From these, the 50 identified by the search engine as most relevant were selected. The 26 listed in Table 2 were those that met all three of the above inclusion criteria. The literature selection process is presented diagrammatically in Figure 2. Each paper was reviewed by two authors (EK and SM) and in cases of disagreement, consensus was required. In what follows we refer to papers listed in Table 2 by their number in column 1.

**Figure 2:**
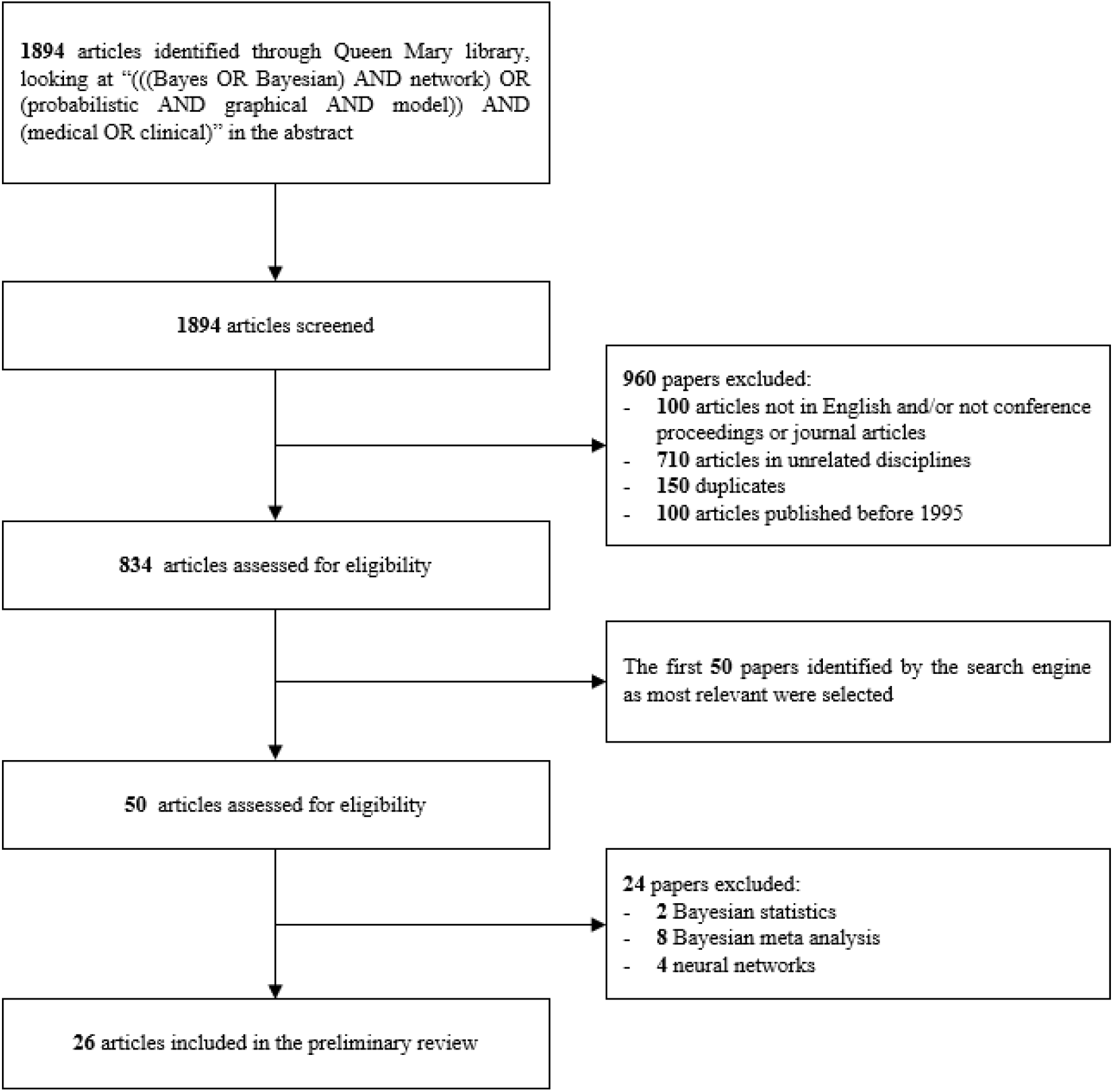
Preliminary review literature selection

**Table 2:**
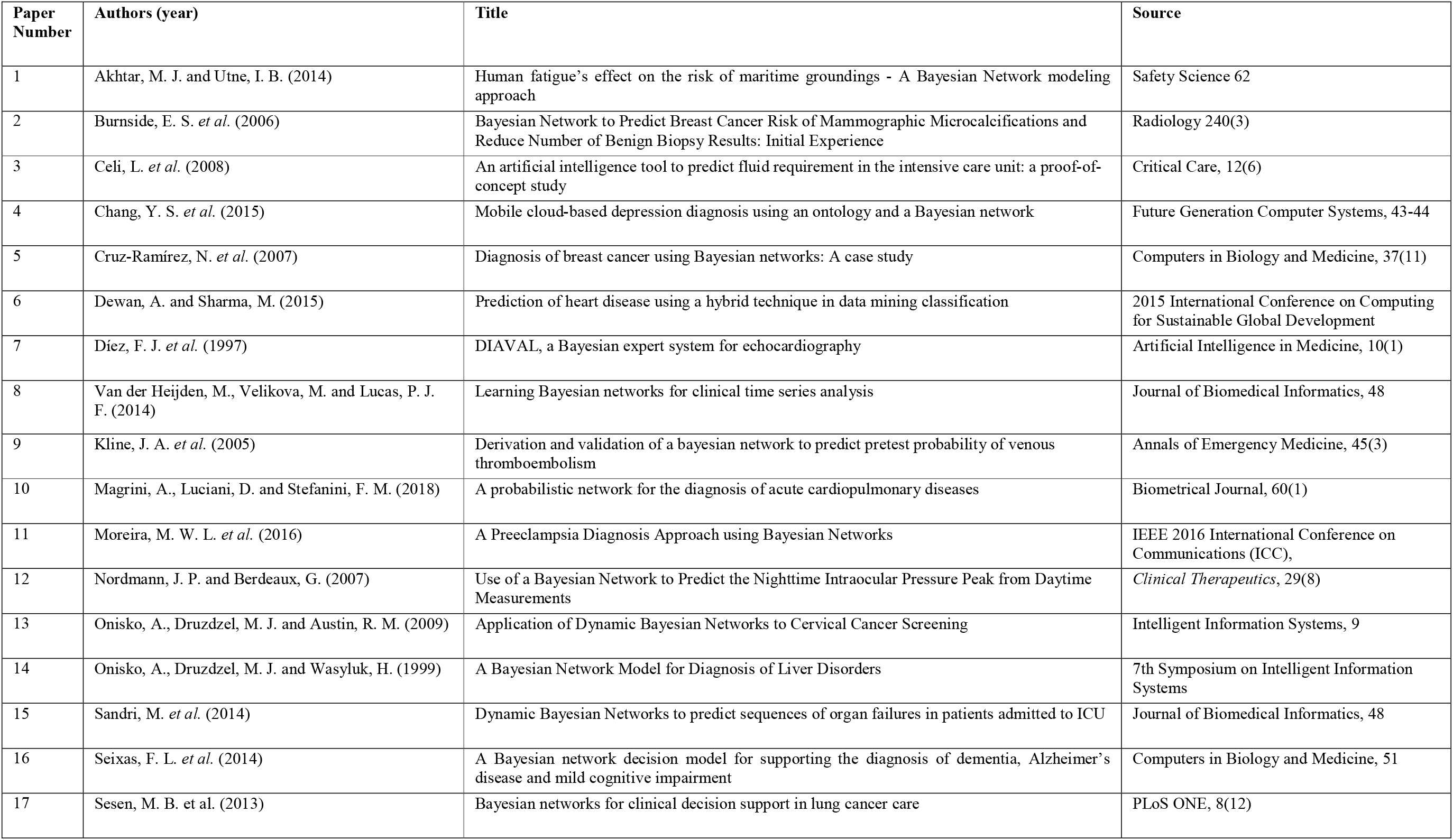
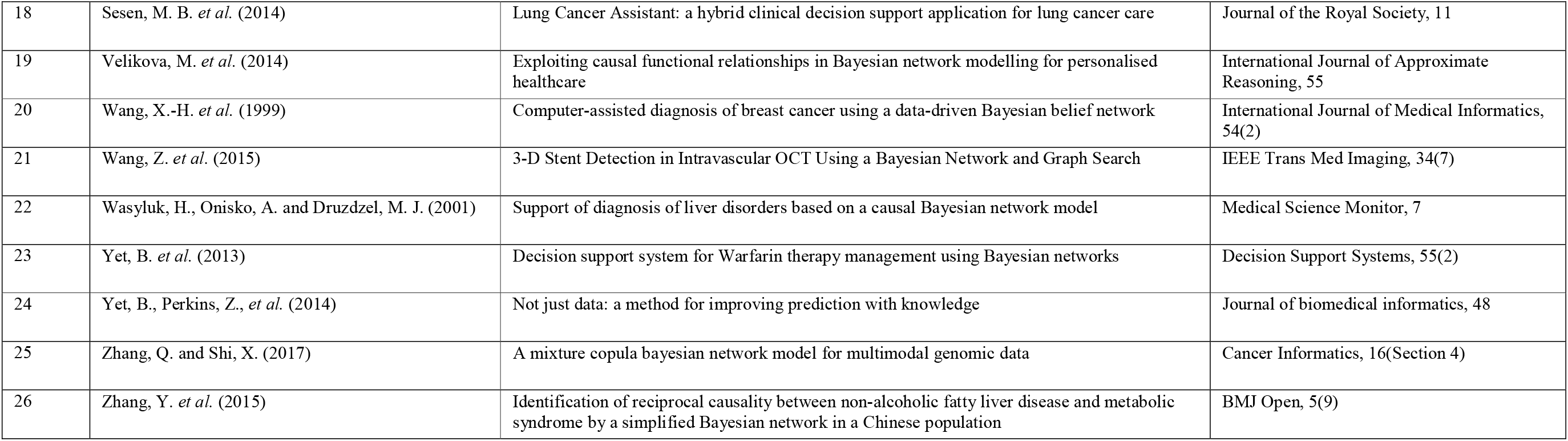
papers in preliminary review that met all inclusion characteristics

### BN development process

Only 5 out of the 26 reviewed papers provided a detailed repeatable *overall development process* [24, 16, 1, 6, 10]. Similarly, from the 13 papers in which the BN structure was elicited from knowledge, only 4 provided a generic structure such as a template [17, 18, 19, 10]. Moreover, no paper provided a detailed explanation of how the knowledge was elicited from experts. Often, references were given to activities such as interviews that had been conducted to elicit the expert knowledge without providing details of the interview process, questions asked, or responses received from the participants. While it is not uncommon to elicit the BN structure from knowledge, eliciting BN parameters from knowledge is generally difficult. In 5 of the 26 reviewed papers, we identified BN models whose parameters were elicited partially or completely from knowledge [2, 23, 19, 11, 10]. From these 5 papers, only one provided a repeatable *parameter elicitation process* [24]. Similar results were also identified where BN structure and/or parameters were learned from data. From the 15 papers where the BN structure was learned from data, only 4 provided detail of the *structure learning process* [17, 18, 8, 16]. BN parameters were learned from data in 24 of the 26 papers. yet only 3 offered detailed description of the *parameter learning process* and the way missing data were treated [23, 8, 16].

### BN adoption in healthcare

The frequency of elements related to BN adoption in healthcare is presented in Figure 3. Most of the reviewed papers clearly described the *clinical benefit* of the medical BN and provided BNs with acceptable *accuracy*. However, *generalisability* was identified only once [24]. Generalisability is important for achieving BN adoption in healthcare as a model could not be used widely in practice if it has not been shown to work on alternative populations. This strongly indicates a research gap limiting the application of BNs to populations that differ from the one used in the original training of the model. Two of the rarest elements to identify were the model’s *usability* and *impact* (either potential or actual). This agrees with our initial remark that there is a significant gap between developing an accurate model and having a useful and usable model that can be adopted and have an impact in healthcare. In this preliminary review no BN adhered to all the rules necessary for having a ‘useful’ model. **Finally, and crucially, no paper in the preliminary review collection provided any indication at all that the BN models were in current clinical use**.

**Figure 3:**
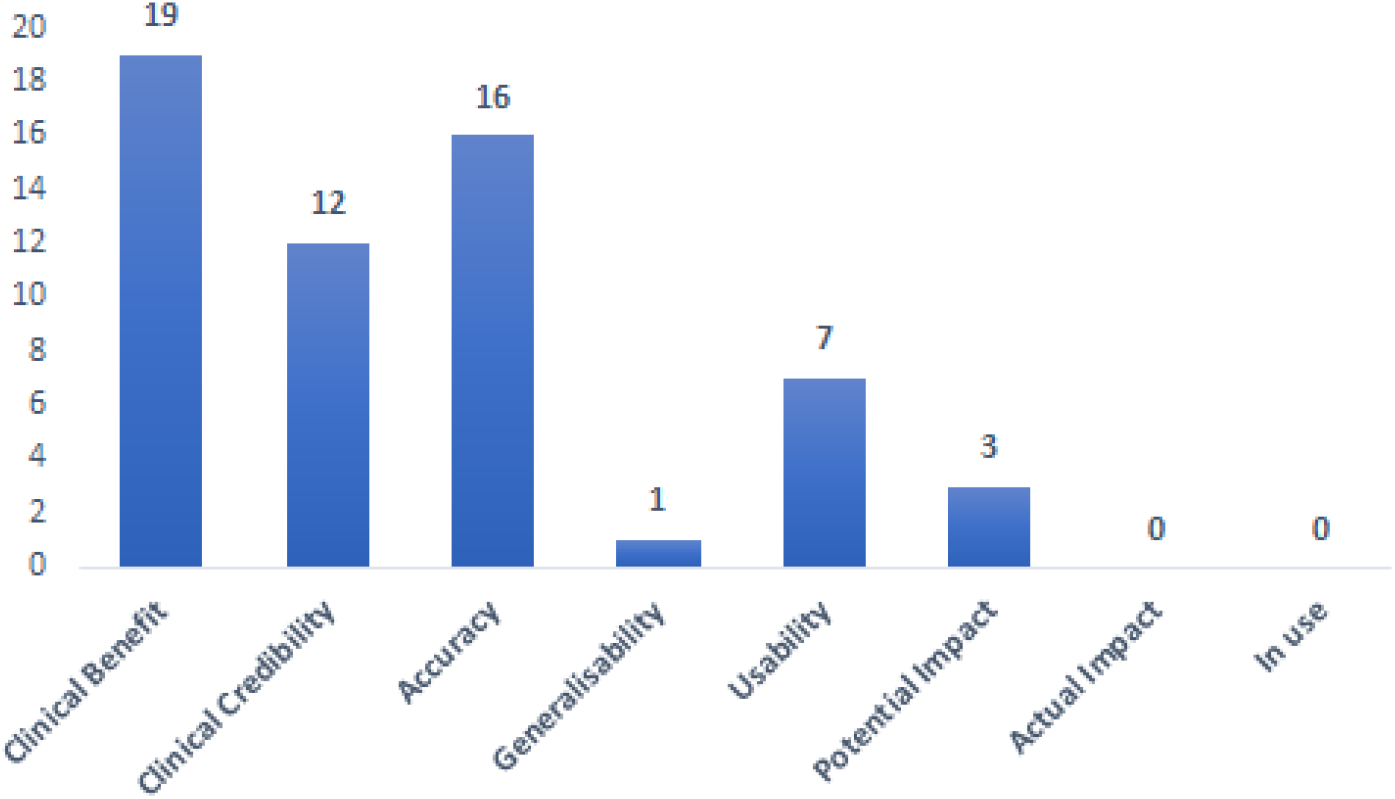
Frequency of the elements related to the BN adoption in healthcare

### 4. Summary and the way forward

This small-scale preliminary review found that literature on medical BNs can be broadly grouped into: (1) method-driven papers, where the focus is on a particular aspect of the modelling methodology; and, (2) case-driven papers, where the focus is on the medical application.

The key findings are that:

1. The literature lacks a well described overall BN development process;
2. Most works only briefly present the way the BN structure and parameters were elicited or learned from knowledge or data, respectively. This makes their methodology difficult, if not impossible, to replicate.
3. Important elements, such as the BN’s generalisability, usability and impact, which are necessary for having a useful model, have received little or no attention. This creates a fundamental gap between developing an accurate BN and adopting it in healthcare.
4. Despite immense interest and research effort in BNs to support clinical decision making, it appears that negligible clinical adoption of *published* BNs into healthcare practice has occurred.

The fact that BNs do not appear to have achieved a significant (publicly accessible) contribution to healthcare suggests that researchers in BNs for healthcare must start to address serious challenges. While our analysis used a sound review framework, it is only a preliminary review where a small number of papers have been selected based on their relevance to the search term to establish the rationale for a more comprehensive investigation. We believe systematic or scoping reviews that summarise and organise the research contributions in the domain may help. The lack of such reviews has likely led to duplication of effort, while allowing important research gaps to stay hidden. The high volume of published works and lack of significant attempts to systematically review the domain are significant motivating factors for undertaking a systematic or scoping review to summarise the literature and highlight research gaps and future directions. A robust scoping review conforming to the PRISMA statement is underway and promises further contributions.

## Data Availability

No data availability

## Contributors

EK prepared the first draft. EK proposed the preliminary framework, which was refined by SM, KD and NF. EK and SM conducted the preliminary review. KD and NF supervised the research. All authors contributed, commented and approved the final draft.

